# Trust in large language model-based solutions in healthcare among people with and without diabetes: a cross-sectional survey from the Health in Central Denmark cohort

**DOI:** 10.1101/2025.02.24.25322734

**Authors:** Jonas R. Schaarup, Anders Aasted Isaksen, Kasper Norman, Lasse Bjerg, Adam Hulman

## Abstract

**Background:** Large language models have gained significant public awareness since ChatGPT’s release in 2022. This study describes the perception of chatbot-assisted healthcare among people with and without diabetes.

**Methods:** In 2024, an online survey was sent to 136,229 people, aged 18–89 years in the Health in Central Denmark cohort, including eight questions related to the perception of AI and chatbots. Questions assessed trust in chatbots in various healthcare scenarios (lifestyle, diagnostic, contact with general practitioner (GP), and emergency contact) alongside participants’ level of experience with ChatGPT. In one item, participants were randomly presented with either a more severe (emergency) or less severe (GP contact) scenario. We used multinomial logistic regression to investigate the association of diabetes status and demographic characteristics with trust in chatbots in different scenarios.

**Findings:** 39,109 participants completed the questionnaire. The majority were aware of AI (94%), though fewer had heard of ChatGPT (76%), and only 21% had tried it. Most participants trusted chatbots with involvement of healthcare professionals (HCP) (49-55%), while few trusted without them (3–6%). The degree of trust depended on the severity of the scenario, demonstrated by lower odds (OR: 0.63 [95% CI: 0.60: 0.66]) of trusting the chatbot under the control of HCP in emergency care compared to contact with the general practitioner. Type 2 diabetes but not type 1 diabetes was associated with less trust in chatbots than people without diabetes. Moreover, age, sex, education, and experience with ChatGPT also had an impact on trust.

**Interpretation:** Chatbots are seen as supportive tools among public users when controlled by HCPs but are met with more skepticism in more severe situations. Digital exclusion risks and demographic differences, such as age, sex, and disease-specific conditions (e.g., type 2 diabetes) needs, must be addressed to ensure equitable and meaningful implementation.

**Research in Context:** *Evidence before this study:* Earlier studies have highlighted the generally positive attitudes of patients and the public towards the applications of artificial intelligence (AI) in healthcare. However, it noted a lack of clear characteristics associated with the acceptance of AI, with many patients preferring AI solutions to remain under human supervision rather than fully replacing healthcare professionals (HCPs). Since ChatGPT emerged in 2022, AI tools have been widely available to the general public, and many healthcare-specific chatbots are now being evaluated in random control trails. Some patients are already turning to tools like ChatGPT for medical advice, further underscoring the need to understand user perceptions, particularly in relation to diabetes and other characteristics, as these technologies may become integrated into care. Our earlier study showed that among AI applications, chatbots were the most controversial when used in emergency care. Thus, understanding the perception of chatbots in different healthcare contexts is needed, as the level of controversy may depend on their specific role in healthcare.

*Added value of this study:* Our study expands on previous work by engaging a larger cohort of 39,109 participants, which includes a comprehensive representation of older adults and individuals with and without diabetes. Our survey was conducted between February-May 2024, a time when ChatGPT had been accessible for more than 1 year. We assessed trust in chatbot-based healthcare solutions, revealing that, while the majority accepted chatbot assistance under human control, individuals with type 2 diabetes exhibited less trust in such applications compared to those without diabetes or type 1 diabetes. Our findings underscore that the severity and acuteness of healthcare scenarios influenced trust levels.

*Implications of all available evidence:* Our findings suggest that while AI and chatbots are becoming widely available, uncertainty about their benefits and risks in healthcare persists. People view healthcare professionals as playing an important role in supporting them, particularly in severe scenarios, toward adopting chatbot solutions. A patient-centered approach is necessary, with tailored solutions to address variations in trust based on factors such as diabetes status, age, sex, and education. Ensuring the involvement of vulnerable populations, such as the elderly and those with type 2 diabetes, is key to avoiding digital exclusion and making chatbot solutions accessible and meaningful.

## Introduction

Chatbots are software applications designed to simulate human-like conversations with users. In recent years, large language model (LLM)-based chatbots such as ChatGPT have gained increasing attention for its potential to revolutionise healthcare domains by handling large volumes of texts (e.g. electronic health records) and providing access to vast knowledge via natural language interactions^1^. Powerful tools like ChatGPT have become accessible to everyone with a computer or smartphone and well-known in the general public^1^. At the same time, demographics are shifting in large parts of the western countries where the populations are aging, resulting in changes in chronic disease burden, as e.g. more people are living with diabetes^2^. Thus, efficient healthcare solutions are required to meet the growing needs of prevention and patient care^3^.

In previous surveys on public perception of AI in healthcare, the use of chatbots in contact with health services was the most polarizing scenario^4,5^. Our previous study showed that people with diabetes were less likely to accept the replacement of healthcare professionals (HCP) with AI solutions compared with people without diabetes^4^. Still, the majority would have accepted AI solutions as long as healthcare providers maintain control, which highlights that human interactions and accountability is important in care with AI^4-6^. Since the release of ChatGPT in 2022, chatbots have become an increasing part of public discourse and policy-making^7^, being used widely in daily life by the public, and even by patients and doctors e.g. to search medical information^8,9^. In 2023, there were registered 57 ongoing clinical trials utilizing chatbots in healthcare^10^. Given the current focus on LLM-based chatbots, there is a need to assess the perception and use of ChatGPT among patients, and their trust in its use in healthcare.

Therefore, we aim to describe the trust in chatbots in healthcare, and how trust differs in various healthcare scenarios. Moreover, we will investigate to which degree trust in chatbots depends on having diabetes along with other characteristics. We hypothesize that people with diabetes, as frequent healthcare users, are more skeptical of chatbots, exhibiting lower trust in them compared to those without diabetes. Moreover, we hypothesise that the degree of trust depends on the acuteness and the severity of the scenario.

## Methods

### Study population

All individuals aged 18-89 years with diabetes (type 1 or type 2), residing in the Central Denmark Region, and a matched sample (matched on age, sex, and municipality) without diabetes were invited to participate in a survey in 2020, forming the Health in Central Denmark (HICD) cohort^11^, with a subgroup followed up in 2022^4^. Diabetes status (type 1, type 2, or without diabetes) was determined using the Danish registries based on a previously described open-sourced algorithm^12^. In 2024, a new survey was conducted, inviting 136,229 participants, providing the basis of our current study.

### Data collection

Participants received a link to an online survey via the Danish Digital Post service or by mail. In addition to our survey on views on AI, chatbot, and data sharing, the questionnaire covered themes such as self-reported health, medication use, dental health, and diet, among others. The cohort was linked to the Danish national registries through the civil personal registration number, a unique identifier assigned to all residents in Denmark.

We adapted our previous pilot-tested questionnaire on AI in general, to focus exclusively on the perception of chatbots^4^. The questionnaire included eight close-ended questions related to use of chatbots, benefit and risk about general AI in healthcare, use of ChatGPT.

Four hypothetical scenarios, where chatbots might assist or replace human clinical care in the future, were presented to all participants (Box 1). All participants were asked about scenarios involving the use of chatbots to support lifestyle recommendations and early detection of diagnoses. To determine whether trust depends on the acuteness of the scenario, two scenarios were randomised among participants. They were assigned to respond to the same scenarios but involving contact to different medical services: a general practitioner or the emergency service (see Fig 1). The four possible answers in all scenarios were ‘Yes’; ‘Yes, but I would also seek information from HCP’ or ‘Yes, but only with control by HCP’ depending on the scenario; ‘No’; ‘I don’t know’. The full questionnaire is available online^13^. The Danish word for “trust” (“at stole på”) refers to having confidence in something, believing it to be reliable, truthful, or capable. We asked participants, “Would you trust this assessment?” to understand their level of confidence and faith in the integrity or abilities of the chatbot in the given scenario. Age, sex, and educational level (<10 years, 10-15 years, >15 years) were extracted from the Danish registries.

**Figure 1:**
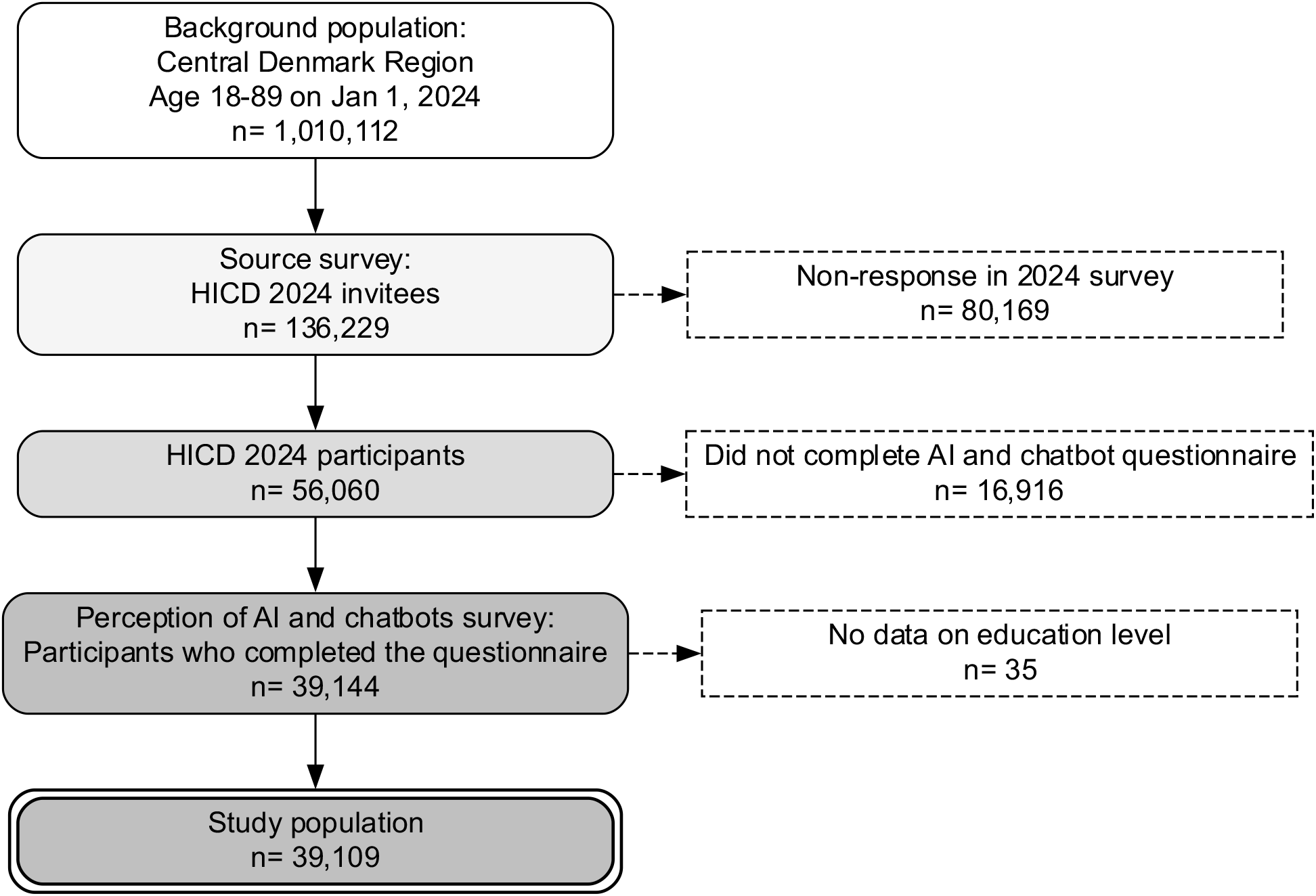
Study flowchart.

#### Box 1

Presented chatbot-based solutions in different healthcare scenarios

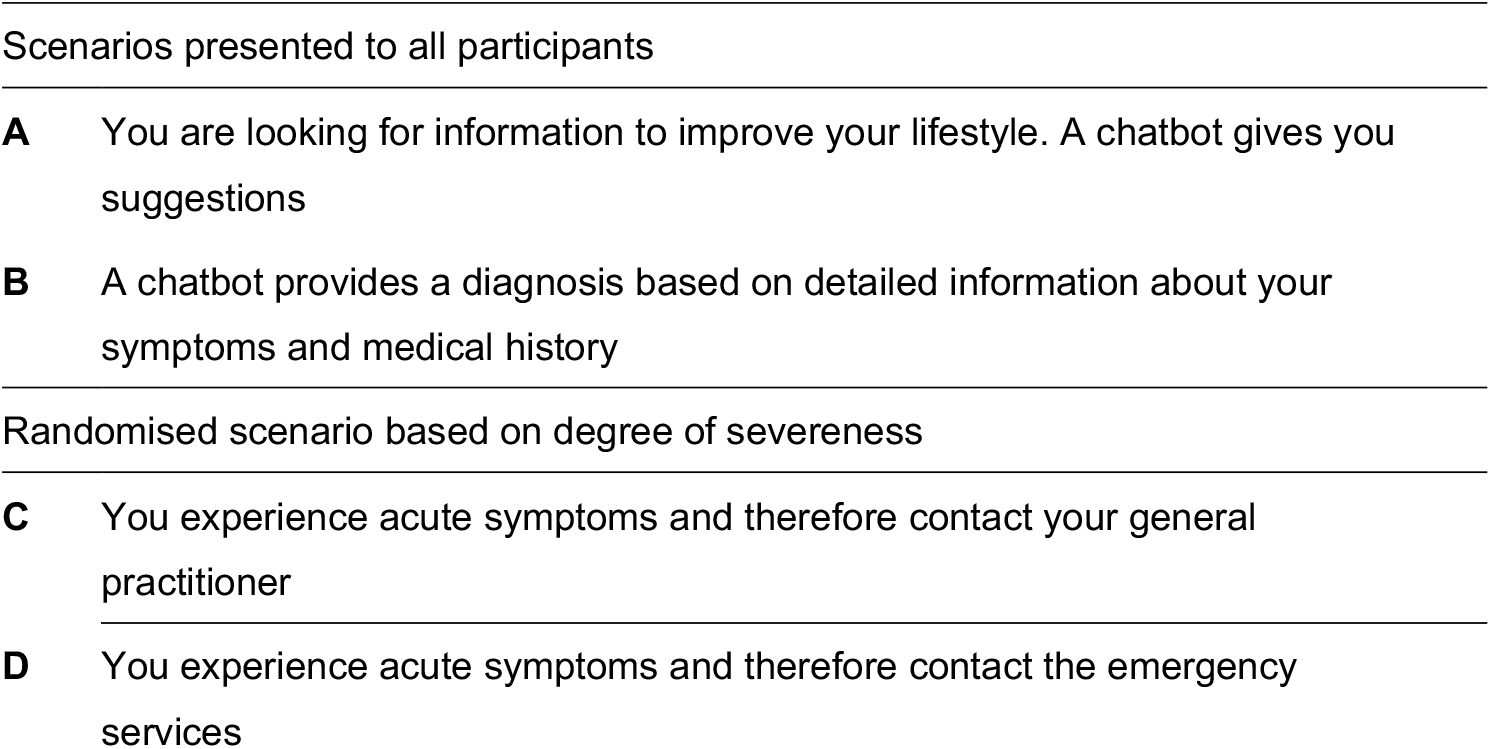

### Statistical analysis

Population characteristics were described by diabetes status. Categorical variables were summarised using frequencies and percentages, while continuous variables were described using the mean and standard deviation (SD).

Distribution of responses to the scenarios were presented as percentages with waffle plots. We used multinomial logistic regression to investigate the association between diabetes status along other characteristics (age, sex, education length) and trust in chatbot in the different scenarios. All determinants were included in the same model. In a separate model, we further included participants’ use and knowledge of ChatGPT as a separate factor. We used multinomial logistic regression to investigate whether the trust of chatbots depended on the acuteness of the contact (questions C and D in Box 1), comparing the effect of randomization to a general practitioner (GP) versus emergency contacts on chatbot trust. The answer “No” was used as the reference category for the outcome.

Complete case analyses were conducted. The statistical analyses were performed using the R statistical computing environment (R Foundation for Statistical Computing, Vienna, Austria; version 4.1) and we used the waffle, nnet and dplyr packages to conduct analyses and for illustrations.

## Results

From the invited population, 39,109 participants (29 % response rate) completed the AI-related questions in the HICD 2024 survey (Figure 1). The mean (SD) age of the respondents was 63 years (SD: 11), with 46% being women. 60% did not have diabetes, 4% had type 1 diabetes, and 36% had type 2 diabetes. Full demographic details and questionnaire responses, shown by diabetes status, are presented in Table 1. A higher proportion of participants had more than 15 years of education (29%) compared to non-responders (20%) (Table S2).

**Table 1:** Demographics, use of ChatGPT, and benefit and risk of AI by diabetes status.

|  | **Overall**<br>N = 39,109 | **Without diabetes**<br>N = 23,355 | **Type 1 diabetes**<br>N = 1,484 | **Type 2 diabetes**<br>N = 14,270 |
| --- | --- | --- | --- | --- |
| Sex |  |  |  |  |
| Men | 21,147 (54%) | 12,013 (51%) | 778 (52%) | 8,356 (59%) |
| Women | 17,962 (46%) | 11,342 (49%) | 706 (48%) | 5,914 (41%) |
| Age (years) | 63 (11) | 63 (12) | 52 (16) | 64 (10) |
| Education duration |  |  |  |  |
| < 10 years | 4,094 (10%) | 1,941 (8%) | 107 (8%) | 2,046 (14%) |
| > 15 years | 11,244 (29%) | 7,731 (33%) | 451 (30%) | 3,062 (21%) |
| 10-15 years | 23,771 (61%) | 13,683 (59%) | 926 (62%) | 9,162 (64%) |
| Have you ever used ChatGPT? |  |  |  |  |
| Yes, I use it often | 1,217 (3%) | 812 (4%) | 85 (6%) | 320 (2%) |
| Yes, I've tried it | 6,949 (18%) | 4,462 (19%) | 357 (24%) | 2,130 (15%) |
| No, but I've heard of it | 21,587 (55%) | 13,272 (57%) | 768 (52%) | 7,547 (53%) |
| No, I haven't heard of it | 9,356 (24%) | 4,809 (21%) | 274 (18%) | 4,273 (30%) |
| Do you think that the benefits of using artificial intelligence in the healthcare sector outweigh the risks? |  |  |  |  |
| Benefits outweigh the risks | 10,413 (27%) | 6,646 (28%) | 429 (29%) | 3,338 (23%) |
| Risks and benefits<br>are equal | 7,128 (18%) | 4,477 (19%) | 277 (19%) | 2,374 (17%) |
| Risks outweigh the<br>benefits | 3,572 (9%) | 2,018 (9%) | 163 (11%) | 1,391 (10%) |
| I don't know, but I<br>have heard of<br>artificial intelligence | 15,810 (40%) | 9,259 (40%) | 542 (37%) | 6,009 (42%) |
| I don't know, and I<br>have never heard<br>of artificial<br>intelligence | 2,186 (6%) | 955 (4%) | 73 (5%) | 1,158 (8%) |

Values are mean (SD) and frequency (%) for continuous and categorical variables, respectively.

### Awareness and use of ChatGPT and AI

Most responders (76%) had heard of ChatGPT, and 8,166 individuals (21%) had tried or used the application. In contrast to participants with type 1 diabetes or without diabetes, larger proportions (30%) of those with type 2 diabetes had not heard about ChatGPT. Overall, 94% had heard about AI in general.

### Acceptance of AI and chatbots in healthcare scenarios

When asked about the potential benefits and risks of AI in healthcare, 46% (N= 17,996) of participants were uncertain whether the advantages would outweigh the risks. Meanwhile, 27% (10,413) believed that the advantages did outweigh the risks, 9% (3,572) responded the risks were greater, and 6% (2,186) had not heard of AI.

Across all scenarios, except for the more severe emergency call scenario (Scenario D), about half (49-55%) would trust chatbots when it was controlled by HCP (**Figure 2**). For both Scenarios A and B (lifestyle recommendations and diagnoses of symptoms based on medical history) more than 50% trusted chatbots with control by HCP. A proportion of 22-42% expressed that they did not trust the decisions made by the chatbot. Only 3-6 % of the responders trusted chatbot without the control of HCP across the scenarios.

**Figure 2.**
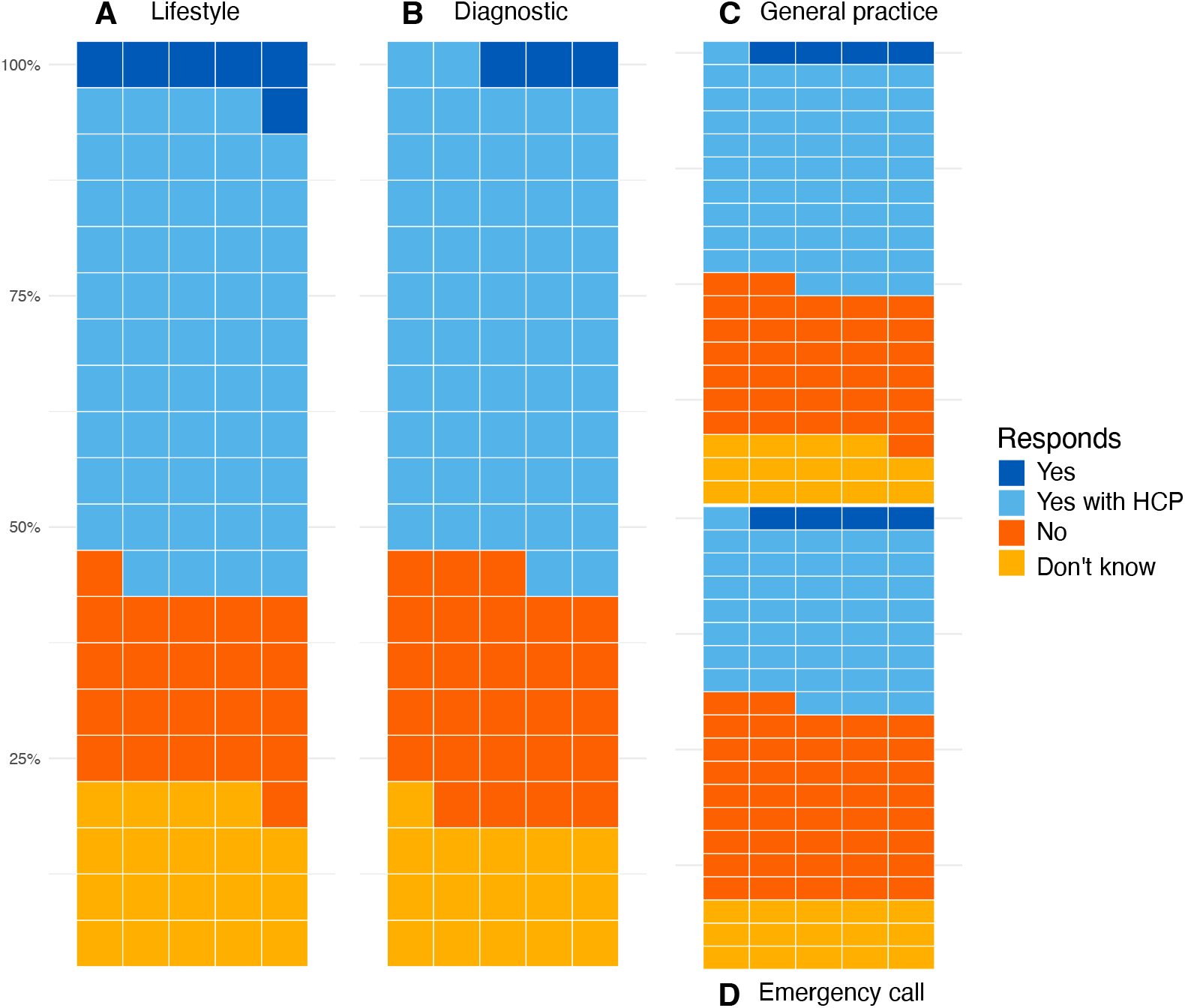
Participants’ views on the use of chatbot-based solutions in different scenarios in healthcare. The answer addresses to which degree the participant ‘would trust the chatbot’. *Percentage distribution on each scenario. Scenario A and B include all participants, while participants were randomized into either scenario C or D. HCP: healthcare professionals*.

The randomisation of less severe (Scenario C) and more severe (Scenario D) scenarios showed that a higher proportion in the less severe scenario trusted chatbots under the control of HCP (less severe: 49% vs. more severe: 39%) (**Figure 2**). Being presented with the more severe scenario, was associated with lower odds (OR: 0.63 [95% CI: 0.60: 0.66]) of trusting the chatbot under the control of HCP as compared to the less severe scenario involving contact with the general practitioner.

The associations between characteristics including diabetes and the trust in chatbot assisting HCP are presented in Figure 3. Odds ratios for other outcome reference categories (trusting AI without HCP, or do not know) are shown in the supplementary materials (**Figure S2**). People with type 2 diabetes were less likely to trust chatbots with control of HCP than responding ‘No’ (OR: 0.74-0.85 for responding trusting the chatbot with control of HCP), while there was no strong evidence for a difference among people with type 1 diabetes OR: 0.91-0.97 (p-value: 0.28-0.70). People with type 1 diabetes were more likely to trust chatbots with HCP when contrasted to those with type-2 diabetes. Adjusting for ChatGPT knowledge and use did not have a major impact on the estimates (**Figure S3)**.

**Figure 3.**
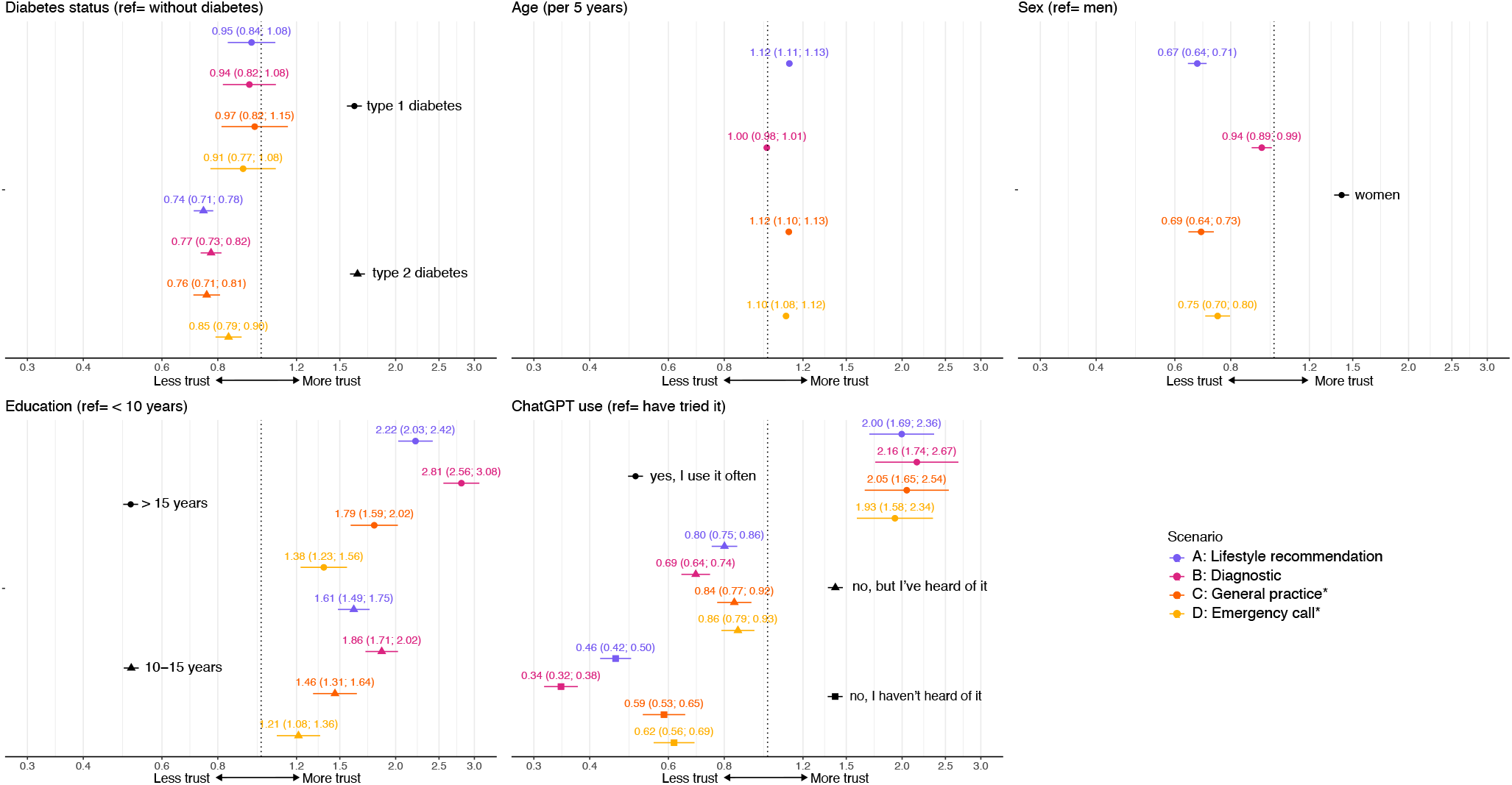
Associations between participant characteristics and trust in chatbot-based technologies (Trust in chatbot with healthcare professionals vs. no trust) in different scenarios in the healthcare setting. *Described by odds ratios (95% CI) from a multinomial logistic regression model adjusted for all displayed variables. Randomized subgroups from either scenario C or D\**.

Women were less likely to trust chatbots with HCP (ORs 0.67-0.94). Except in the scenario of chatbot-assisted diagnosis OR: 1.00 (CI: 0.98 1.01), older age was associated with trusting chatbot in healthcare with control of HCP (OR: 1.10-1.12 per 5 years), but older individuals were also more likely to respond don’t know (**Figure S2**). With increasing age, higher proportions of participants were more likely to be uncertain. After age 30, the trend of trusting chatbots with HCP either remained unchanged or increased across scenarios until age 70+ (**Figure S1**). When contrasting trust in chatbots with HCP against chatbots without HCP, older individuals were less likely to trust chatbots without HCP (**Figure S4**). The duration of education and more experience with ChatGPT was gradually associated with trusting AI with control of HCP compared to not trusting chatbots (**Figure 3)**. Being woman, older age, and shorter education and less experience with ChatGPT was associated with being more likely to report ‘don’t know’, compared to not trusting (**Figure S2**).

## Discussion

In this large cross-sectional study, we investigated the trust in chatbots among people with and without diabetes, spanning a wide age range. Almost all participants have heard about AI, but one in four has not heard of ChatGPT. Most were uncertain whether the benefits of AI in healthcare outweighed its risks. Our findings showed a predominant trust in chatbots for the healthcare system as long as HCP remain part of the process, though some participants expressed uncertainty or lack of trust. Trust in chatbot-assisted services was dependent on the acuteness and severity of the different scenarios, with lower levels of trust observed in more acute health contexts, such as emergency care contacts. Across all scenarios, few participants trusted chatbots without the involvement of HCPs. While participants with type 2 diabetes had lower trust than without diabetes, we did not find any evidence for a difference between people with type 1 diabetes and without diabetes.

Our results suggest that public awareness of AI has increased over recent years, gaining broader public exposure compared to earlier studies^4,14^. Almost half were uncertain about whether AI would benefit healthcare, while 1 in 4 believed the benefits would outweigh the risks. Compared to our earlier study and a study from the UK, these findings suggest greater public uncertainty regarding AI in healthcare^4,14^. In the current study, ChatGPT remains unknown for a fourth of the population, with one out of five having tried it, a figure lower than a recent report from the UK^15^. People who have used chatbots were more open to trust them, although most remain convinced that chatbot solutions could not stand alone and should be used with control, follow-up consultation or second opinion by HCP. This was the common opinion from all participants regardless of diabetes status, except in the scenario of emergency service where the majority would not trust the chatbot. A recent study has exemplified how chatbot-based solutions can be used in clinical settings with control of HCP, in diagnostic aid of patients^16^. Although no evidence was found of improved physician diagnostic performance, there is a notion that chatbots, when operating independently, have occasionally outperformed humans^16^. However, another study showed that ChatGPT performed worse in a comprehensive evaluation when it was used in complex primary care settings^17^. Moreover, chatbots’ potential as a patient and clinician support tool (e.g. medical note-taking, consultations) has been widely recognized, though some risks (e.g. hallucinations and biases) represent concerns^18,19^. Previous studies have highlighted the importance of providing patients with explanations for clinical decisions and ensuring accountability in the decision-making process^20,21^. Thus, strategies for the integration of HCP in chatbot-based solutions remain important.

Despite high trust in chatbots with control of HCP, almost a half of participants either did not trust or were unsure about trusting chatbots. The complexity and perceived importance of the tasks (e.g., lifestyle advice vs. emergency decision-making) seemed to influence public trust. Additionally, patient characteristics such as sex, age, education, and familiarity with ChatGPT impacted acceptance. The trends among factors observed align with findings from a multinational study on perceptions of AI in broader contexts^22^. AI in healthcare, while offering transformative potential, can exacerbate digital exclusion when individuals are unable to access or utilize AI-driven technologies due to barriers. Digital exclusion has been shown to depend on experience with digital technology and socio-demographic factors, emphasizing the need to address barriers and facilitators in AI systems in healthcare^23^. Ignoring user characteristics could lead to digital exclusion which risks differential uptake of chatbot-based solutions and potentially lead to inequalities in access to these healthcare solutions.

In this study, we found that trust in chatbots increased with age when HCPs were involved. However, older adults also showed more “don’t know” responses, reflecting higher uncertainty by age. Thus, older adults are open to using chatbots with HCP support, but some may still lack confidence or familiarity with the technology. Our earlier HICD 2022 study showed that older adults were less likely to accept AI without HCP involvement compared to AI with HCP support. Using the same contrast, in current study, we confirmed these findings.

These uncertainties among age groups highlights the risk of digital exclusion, which may arise from limited digital literacy, access to technology, or hesitation to engage with unfamiliar tools^24^. Nonetheless, a study has shown that older adults are viewing digital health solution as a useful tool^25^, but wants HCPs to be involved.

The use of chatbots in healthcare show potential to empower patients by providing individualized, easily accessible, and timely support with communication and knowledge sharing^8^. Employees at a large Danish diabetes center performed only slightly better than random chance (a coin toss) in distinguishing between chatbot-generated and human-written responses to questions related to diabetes care^26^. Chatbot responses have been found to demonstrate greater empathy compared to doctors’ replies on a social media forum^27^, but patients have shown to be more distrustful of the non-human responses^28^. Having type 2 diabetes was associated with being less likely to trust chatbots compared to people without diabetes. Managing diabetes requires personalized attention to lifestyle, medication, and the monitoring of symptoms related to diabetes complications, which constitutes living with a higher overall disease risk^29^. Such conditions can lead to increased distress and insecurity^29^ demanding human support and guidance, and thus making deviation from usual care less likely. For people with type 1 diabetes, adoption of advanced technologies such as continuous glucose monitors and insulin pumps has become a standard part of disease management over the last decade ^30^. Nearly a third of people with type 2 diabetes reported not having heard of ChatGPT compared to a fifth in people with type 1 diabetes or without diabetes. In our previous study, we found individuals with type 1 diabetes to be less open to AI solutions than people without diabetes, however an emergency contact chatbot scenario was inconclusive and the most controversial of the scenarios studied^4^. In the current study, people with type 2 diabetes were older, which is associated with lower digital literacy and, consequently, higher levels of uncertainty and lower trust in technologies. Adjusting for age and familiarity with ChatGPT did not explain the differences within the diabetes group. Therefore, people with type 1 diabetes appear as open to chatbot solutions as those without diabetes, requiring further investigation into their readiness and trust compared to individuals with type 2 diabetes.

Sex differences in healthcare utilization remain a challenge, with men often lagging in seeking care. In Denmark, women make more use of primary care than men, while men have a higher use of secondary care^31^. As men showed to be more likely to trust chatbot solutions and tend to have higher digital literacy^32^, one hypothesis could be that chatbots might help initiate earlier healthcare contact. On the other hand, studies have shown that women value their interaction and relationship with their healthcare provider more than men^33^. With the potential of chatbot solutions, sex differences should be considered to ensure that chatbots complement and enhance healthcare solutions while not replacing essential aspects of care. Chatbots could serve as an entry point to the healthcare system, potentially encouraging more men to seek medical contact.

Our previous study based on the 2022 questionnaire included a subpopulation of 8,420 participants, who responded to questionnaires about scenarios involving AI in healthcare^4^. Compared to our current findings, this group demonstrated a higher acceptance of chatbots without HCP involvement (10%) than participants in the current study (3-6%). Chatbots remain the most controversial application of AI in healthcare compared to other applications e.g. diagnosis based on images and monitoring by wearable devices. Our findings indicate that public concern about the role of AI in the healthcare system has increased, alongside growing uncertainty about whether the benefits of AI outweigh its risks. It is important to note that the 2022 HICD investigation involved a smaller non-random subset of participants and presented scenarios where AI applications were described as being as good as or better than usual care. In contrast, the current study excluded such explicit assurances, reflecting the rapid acceleration of AI and aligning more closely with the realistic implementation of these scenarios. As such, the differences between the populations and scenarios make direct comparisons challenging.

This study is based on a large population from Central Region Denmark including people with and without diabetes. Such sample size allowed us to conduct randomization of questions to demonstrate impact of severity of scenarios on perception of chatbot-based solutions. Moreover, we have a broad representation of demographic groups by age, sex, education, across diabetes status. The response rate was 29%, therefore certain subgroups are less represented in the questionnaires e.g. people with shorter education who usually are underrepresented. Chatbots are often being updated, affecting the user experience. Thus, we acknowledge our findings are snapshots of perception of chatbots. Moreover, we acknowledge that AI is frequently discussed in the media^34^, highlighting both the opportunities and risks of the technology, which might have influenced participants’ responses. Our study represents a population of predominantly Nordic European descent with access to high-quality diabetes care, and therefore their opinion may not generalise to other populations.

User involvement with groups of diverse representation i.e. age, sex, disease-specific conditions (e.g. diabetes), educational level, digital literacy, ethnic background are needed to understand how AI can be used in a structured and meaningful way in different healthcare solutions both with and without HCP. Patients already use chatbots for self-diagnosis and health recommendations^8^. As the available chatbots are not yet implemented for healthcare purposes in Denmark, there is an urgent need for medically specialized chatbots to prevent unnecessary harm or concerns from current biases in available chatbots. While randomized control trials remains vital to demonstrate safety and effectiveness of the use of chatbots compared to usual care^35^, we emphasise the importance of addressing user concerns to meaningfully integrate chatbots into high-stakes healthcare settings. This could be addressed using qualitative methods.

## Conclusion

Trust in chatbots is highest when controlled by HCP, highlighting their essential role in guiding chatbot adoption. Moreover, trust varies by disease status, for example, people with type 2 diabetes were less trusting of chatbot-based healthcare solutions. This suggests that chatbots should complement rather than replace essential aspects of care, particularly for managing chronic conditions. The degree of trust is also influenced by the severity and acuteness of the health scenario, with greater openness to chatbot-based solutions in less severe settings, such as lifestyle recommendations, highlighting easier opportunities for implementation. Experience with ChatGPT along demographic characteristics was linked to trust. User involvement from patient groups is needed to understand population-specific perceptions and tailor meaningful implementation of chatbot-based interventions in healthcare.

## Acknowledgements

The authors are grateful to all participants of the HICD cohort for their contributions to the study. We would like to express our gratitude to the user panel (‘StenoPanelet’) at Steno Diabetes Center Aarhus for discussing our previous findings from the AI perception study in 2022 and inspiring us to develop the second questionnaire.

## Contributions

KN and LB designed the HICD cohort. JRS, AAI, KN contributed to data management. JRS, AAI, LB, and AH designed the current study and the questionnaire on AI with contributions from all other authors. JRS, AAI and AH conducted the statistical analysis and created visualizations of the results. JRS drafted the initial version of the manuscript. All authors contributed to data interpretation and to the final revisions of the manuscript. All authors agreed to the submission of the manuscript. JRS is the guarantor of this work and, as such, had full access to all data in the study.

## Data availability

The HICD cohort is managed by a steering committee at Steno Diabetes Center, Denmark. The committee encourages interested researchers to use this resource. Details about the data application process can be found on the HICD website (www.hicd.rm.dk). For further inquiries, please contact KN or LB.

## Ethical approval

The HICD study is based on questionnaires and data from the registries, and such research projects do not require ethical approval in Denmark. The HICD cohort is registered in the Central Denmark Region internal register of research projects (reg. no. 1-16-02-165-20).

