## Supplemental material for "Trust in large language model-based solutions in healthcare among people with and without diabetes: a cross-sectional survey from the Health in Central Denmark cohort"

**Table S1:** Distribution by randomization in scenario C and D

| | C: General practitioner<br>N= 19.422 | D: Emergency care<br>N= 19.687 |
|---|---|---|
| Sex | | |
| Men | 10,503 (54%) | 10,644 (54%) |
| Women | 8,919 (46%) | 9,043 (46%) |
| Age (years) | 65 (56, 71) | 65 (56, 71) |
| Education level | | |
| < 10 years | 2,040 (11%) | 2,054 (10%) |
| 10-15 years | 5,605 (29%) | 5,639 (29%) |
| > 15 years | 11,777 (61%) | 11,994 (61%) |
| Diabetes status | | |
| Without diabetes | 11,546 (59%) | 11,809 (60%) |
| Type-1 diabetes | 736 (3.8%) | 748 (3.8%) |
| Type-2 diabetes | 7,140 (37%) | 7,130 (36%) |
| Have you ever used ChatGPT? | | |
| Yes, I use it often | 625 (3.2%) | 592 (3.0%) |
| Yes, I've tried it | 3,440 (18%) | 3,509 (18%) |
| No, but I've heard of it | 10,750 (55%) | 10,837 (55%) |
| No, I haven't heard of it | 4,607 (24%) | 4,749 (24%) |
| Do you think that the benefits of using artificial intelligence in the healthcare sector outweigh the risks? | | |
| Benefits outweigh the risks | 5,200 (27%) | 5,213 (26%) |
| Risks and benefits are equal | 3,578 (18%) | 3,550 (18%) |
| Risks outweigh the benefits | 1,780 (9.2%) | 1,792 (9.1%) |
| I don't know, but I have heard of artificial intelligence | 7,755 (40%) | 8,055 (41%) |
| I don't know, and I have never heard of artificial intelligence | 1,109 (5.7%) | 1,077 (5.5%) |

**Table S2:** Responders characteristics compared to non-responders and background population

| | Background population<br>N =1,010,112 | Invited<br>N= 136,229 | Non-responder<br>N= 97,120 | Responder<br>N= 39,109 |
|---|---|---|---|---|
| Sex | | | | |
| Men | 506,779 (50%) | 74,913 (55%) | 53,766 (55%) | 21,147 (54%) |
| Women | 503,333 (50%) | 61,316 (45%) | 43,354 (45%) | 17,962 (46%) |
| Age (years) | 47 (31, 61) | 62 (52, 70) | 61 (50, 70) | 65 (56, 71) |
| Education level | | | | |
| < 10 years | 111,438 (11%) | 21,402 (16%) | 17,308 (18%) | 4,094 (10%) |
| 10-15 years | 584,258 (60%) | 81,863 (61%) | 58,092 (61%) | 23,771 (61%) |
| > 15 years | 285,626 (29%) | 30,367 (23%) | 19,123 (20%) | 11,244 (29%) |
| Diabetes status | | | | |
| Without diabetes | 945,310 (94%) | 77,814 (57%) | 54,459 (56%) | 23,355 (60%) |
| Type-1 diabetes | 5,913 (0.6%) | 5,698 (4.2%) | 4,214 (4.3%) | 1,484 (3.8%) |
| Type-2 diabetes | 58,889 (5.8%) | 52,717 (39%) | 38,447 (40%) | 14,270 (36%) |

**Table S3**: The effect of randomization on trust in chatbots for emergency contacts compared to trust in chatbots for general practitioners

| Respond | OR | conf.low | conf.high |
|---|---|---|---|
| Yes | 0.81 | 0.73 | 0.91 |
| Yes, but only with control by healthcare professionals | 0.63 | 0.60 | 0.66 |
| Don't know | 0.81 | 0.76 | 0.86 |
| No (ref.) | 1.00 | 1.00 | 1.00 |

**Table S4:** Main results from Figure 3 of diabetes status with reference category changed to type 2 diabetes

| Scenario | Group | Responds | Odds ratio | conf.low | conf.high | p.value |
|---|---|---|---|---|---|---|
| Lifestyle | Without diabetes | Yes | 1.1992530 | 1.0540735 | 1.364428 | 0.006 |
| Lifestyle | Type-1 diabetes | Yes | 1.1251524 | 0.8214024 | 1.541228 | 0.463 |
| Lifestyle | Without diabetes | Yes, but I will also seek additional information, for example, from healthcare | 1.3457125 | 1.2796359 | 1.415201 | 0.000 |
| Lifestyle | Type-1 diabetes | Yes, but I will also seek additional information, for example, from healthcare | 1.2810247 | 1.1300621 | 1.452154 | 0.000 |
| Lifestyle | Without diabetes | Don't know | 1.0671222 | 0.9984067 | 1.140567 | 0.056 |
| Lifestyle | Type-1 diabetes | Don't know | 1.0388310 | 0.8563011 | 1.260269 | 0.699 |
| Diagnostic | Without diabetes | Yes | 1.3464083 | 1.2210799 | 1.484600 | 0.000 |
| Diagnostic | Type-1 diabetes | Yes | 1.1211745 | 0.8904026 | 1.411757 | 0.331 |
| Diagnostic | Without diabetes | Yes, but only with follow-up consultations with healthcare professionals. | 1.2942405 | 1.2267712 | 1.365420 | 0.000 |
| Diagnostic | Type-1 diabetes | Yes, but only with follow-up consultations with healthcare professionals. | 1.2182435 | 1.0601907 | 1.399859 | 0.005 |
| Diagnostic | Without diabetes | Don't know | 1.0832365 | 1.0145884 | 1.156529 | 0.017 |
| Diagnostic | Type-1 diabetes | Don't know | 1.1455236 | 0.9518228 | 1.378644 | 0.151 |
| Emergency care | Without diabetes | Yes | 1.1442464 | 0.9706821 | 1.348845 | 0.108 |
| Emergency care | Type-1 diabetes | Yes | 1.4304686 | 0.9859978 | 2.075299 | 0.059 |
| Emergency care | Without diabetes | Yes, but only with control by healthcare professionals | 1.1826348 | 1.1058759 | 1.264722 | 0.000 |
| Emergency care | Type-1 diabetes | Yes, but only with control by healthcare professionals | 1.0778033 | 0.9070481 | 1.280704 | 0.395 |
| Emergency care | Without diabetes | Don't know | 1.0124047 | 0.9262734 | 1.106545 | 0.786 |

| Scenario | Group | Responds | Odds ratio | conf.low | conf.high | p.value |
|---|---|---|---|---|---|---|
| Emergency care | Type-1 diabetes | Don’t know | 1.0771490 | 0.8407282 | 1.380054 | 0.557 |
| General practitioner | Without diabetes | Yes | 0.9826092 | 0.8317477 | 1.160834 | 0.837 |
| General practitioner | Type-1 diabetes | Yes | 1.2772647 | 0.8741062 | 1.866370 | 0.206 |
| General practitioner | Without diabetes | Yes, but only with control by healthcare professionals | 1.3234415 | 1.2357715 | 1.417331 | 0.000 |
| General practitioner | Type-1 diabetes | Yes, but only with control by healthcare professionals | 1.2803257 | 1.0747472 | 1.525227 | 0.006 |
| General practitioner | Without diabetes | Don’t know | 1.0143038 | 0.9242918 | 1.113082 | 0.765 |
| General practitioner | Type-1 diabetes | Don’t know | 1.1799120 | 0.9071274 | 1.534726 | 0.217 |

**Figure S1:** Participants' views on the use of chatbot-based solutions in different scenarios in healthcare by age groups. The answer addresses to which degree the participant 'would trust the chatbot'.

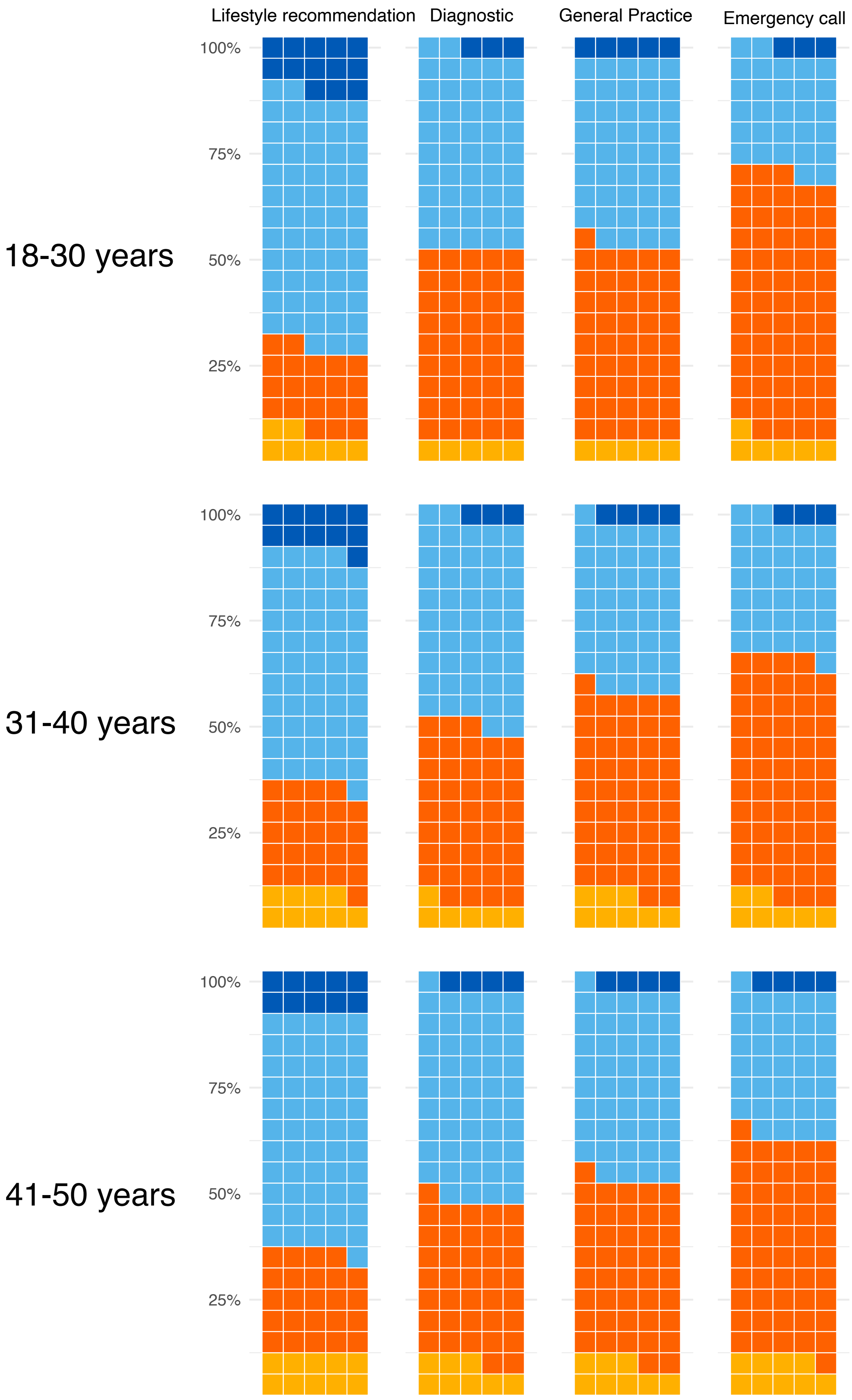

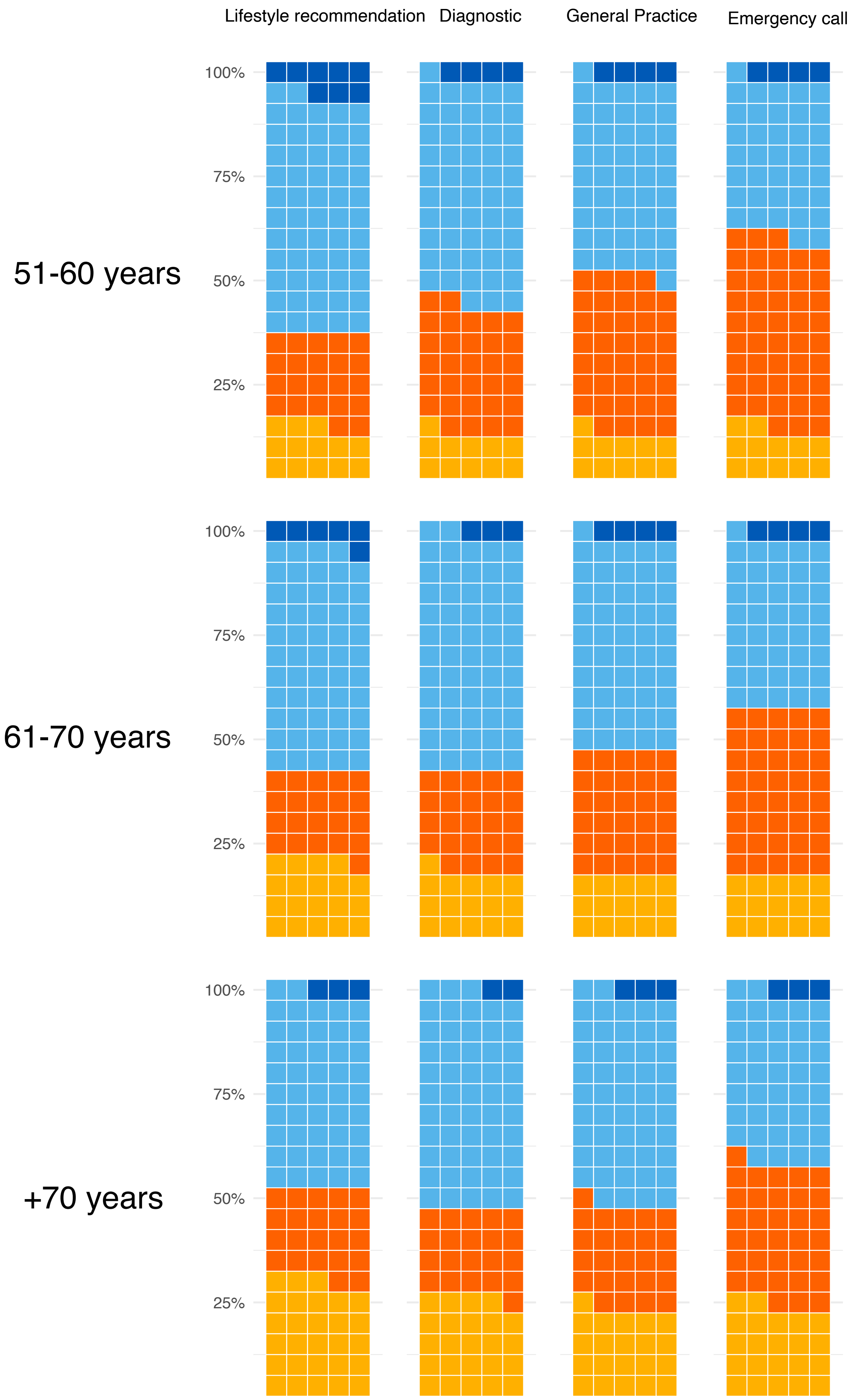

Lifestyle recommendation
Diagnostic
General Practice
Emergency call
51-60 years
61-70 years
+70 years
100%
75%
50%
25%

**Figure S2:** Associations between participants characteristics and a positive response to chatbot-based technologies (reference: No trust) in different scenarios in the healthcare setting.

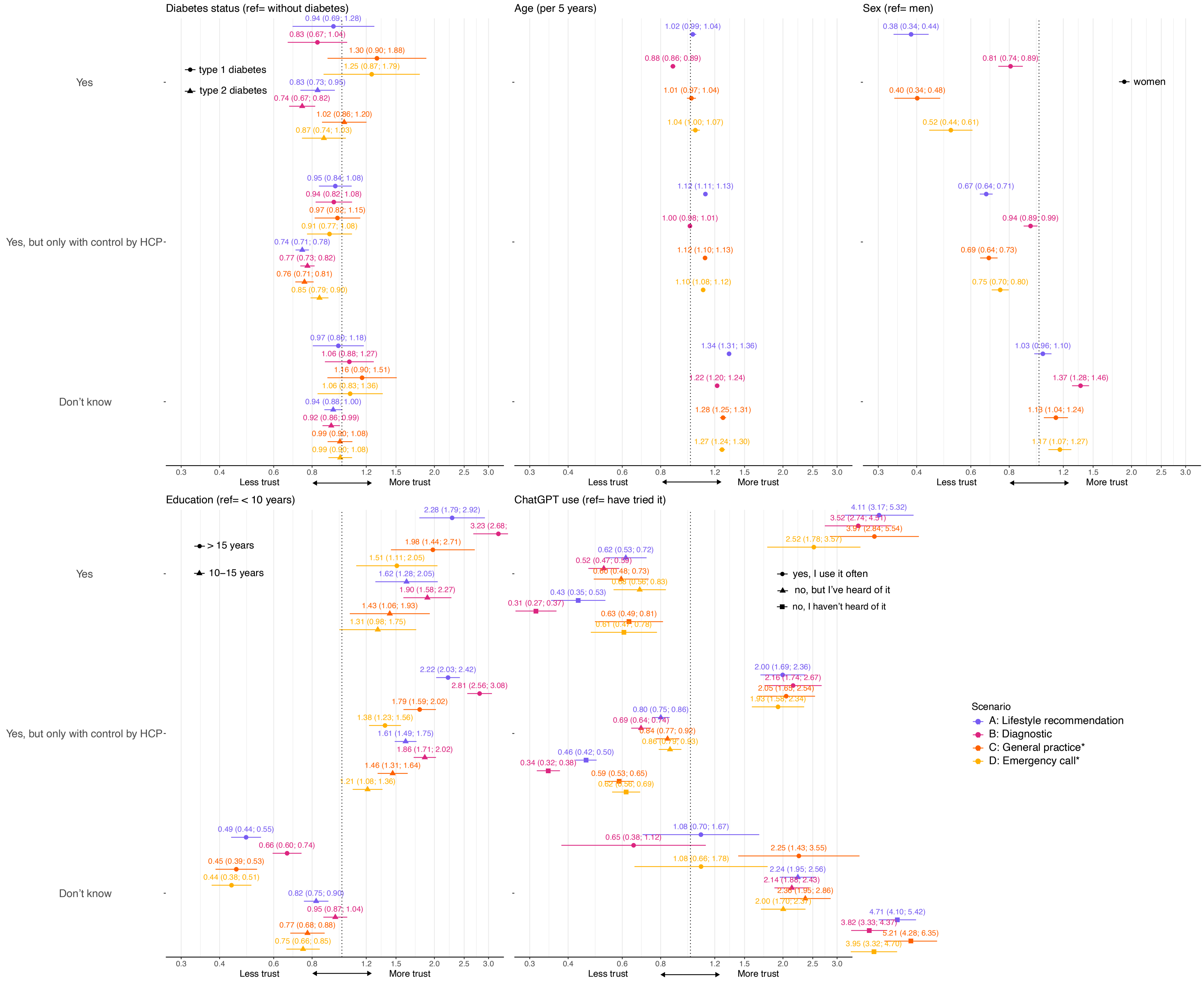
Diabetes status (ref= without diabetes)
Age (per 5 years)
Sex (ref= men)
Education (ref= < 10 years)
ChatGPT use (ref= have tried it)
Yes
Yes, but only with control by HCP
Don't know
type 1 diabetes
type 2 diabetes
women
> 15 years
10–15 years
yes, I use it often
no, but I've heard of it
no, I haven't heard of it
Scenario
A: Lifestyle recommendation
B: Diagnostic
C: General practice*
D: Emergency call*
0.3 0.4 0.6 0.8 1.2 1.5 2.0 2.5 3.0
Less trust
More trust
0.94 (0.69; 1.28)
0.83 (0.67; 1.04)
1.30 (0.90; 1.88)
1.25 (0.87; 1.79)
0.83 (0.73; 0.95)
0.74 (0.67; 0.82)
1.02 (0.86; 1.20)
0.87 (0.74; 1.03)
0.95 (0.84; 1.08)
0.94 (0.82; 1.08)
0.97 (0.82; 1.15)
0.91 (0.77; 1.08)
0.74 (0.71; 0.78)
0.77 (0.73; 0.82)
0.76 (0.71; 0.81)
0.85 (0.79; 0.90)
0.97 (0.80; 1.18)
1.06 (0.88; 1.27)
1.16 (0.90; 1.51)
1.06 (0.83; 1.36)
0.94 (0.88; 1.00)
0.92 (0.86; 0.99)
0.99 (0.90; 1.08)
0.99 (0.90; 1.08)
1.02 (0.99; 1.04)
0.88 (0.86; 0.89)
1.01 (0.97; 1.04)
1.04 (1.00; 1.07)
1.12 (1.11; 1.13)
1.00 (0.98; 1.01)
1.12 (1.10; 1.13)
1.10 (1.08; 1.12)
1.34 (1.31; 1.36)
1.22 (1.20; 1.24)
1.28 (1.25; 1.31)
1.27 (1.24; 1.30)
0.38 (0.34; 0.44)
0.81 (0.74; 0.89)
0.40 (0.34; 0.48)
0.52 (0.44; 0.61)
0.67 (0.64; 0.71)
0.94 (0.89; 0.99)
0.69 (0.64; 0.73)
0.75 (0.70; 0.80)
1.03 (0.96; 1.10)
1.37 (1.28; 1.46)
1.13 (1.04; 1.24)
1.17 (1.07; 1.27)
2.28 (1.79; 2.92)
3.23 (2.68;
1.98 (1.44; 2.71)
1.51 (1.11; 2.05)
1.62 (1.28; 2.05)
1.90 (1.58; 2.27)
1.43 (1.06; 1.93)
1.31 (0.98; 1.75)
2.22 (2.03; 2.42)
2.81 (2.56; 3.08)
1.79 (1.59; 2.02)
1.38 (1.23; 1.56)
1.61 (1.49; 1.75)
1.86 (1.71; 2.02)
1.46 (1.31; 1.64)
1.21 (1.08; 1.36)
0.49 (0.44; 0.55)
0.66 (0.60; 0.74)
0.45 (0.39; 0.53)
0.44 (0.38; 0.51)
0.82 (0.75; 0.90)
0.95 (0.87; 1.04)
0.77 (0.68; 0.88)
0.75 (0.66; 0.85)
4.11 (3.17; 5.32)
3.52 (2.74; 4.51)
3.97 (2.84; 5.54)
2.52 (1.78; 3.57)
0.62 (0.53; 0.72)
0.52 (0.47; 0.59)
0.60 (0.48; 0.73)
0.68 (0.56; 0.83)
0.43 (0.35; 0.53)
0.31 (0.27; 0.37)
0.63 (0.49; 0.81)
0.61 (0.47; 0.78)
2.00 (1.69; 2.36)
2.16 (1.74; 2.67)
2.05 (1.65; 2.54)
1.93 (1.58; 2.34)
0.80 (0.75; 0.86)
0.69 (0.64; 0.74)
0.84 (0.77; 0.92)
0.86 (0.79; 0.93)
0.46 (0.42; 0.50)
0.34 (0.32; 0.38)
0.59 (0.53; 0.65)
0.62 (0.56; 0.69)
1.08 (0.70; 1.67)
0.65 (0.38; 1.12)
2.25 (1.43; 3.55)
1.08 (0.66; 1.78)
2.24 (1.95; 2.56)
2.14 (1.88; 2.43)
2.36 (1.95; 2.86)
2.00 (1.70; 2.37)
4.71 (4.10; 5.42)
3.82 (3.33; 4.37)
5.21 (4.28; 6.35)
3.95 (3.32; 4.70)

**Figure S3:** Associations between participants characteristics and a positive response to chatbot-based technologies (reference: No trust) in different scenarios in the healthcare setting. All estimates are adjusted for use of ChatGPT.

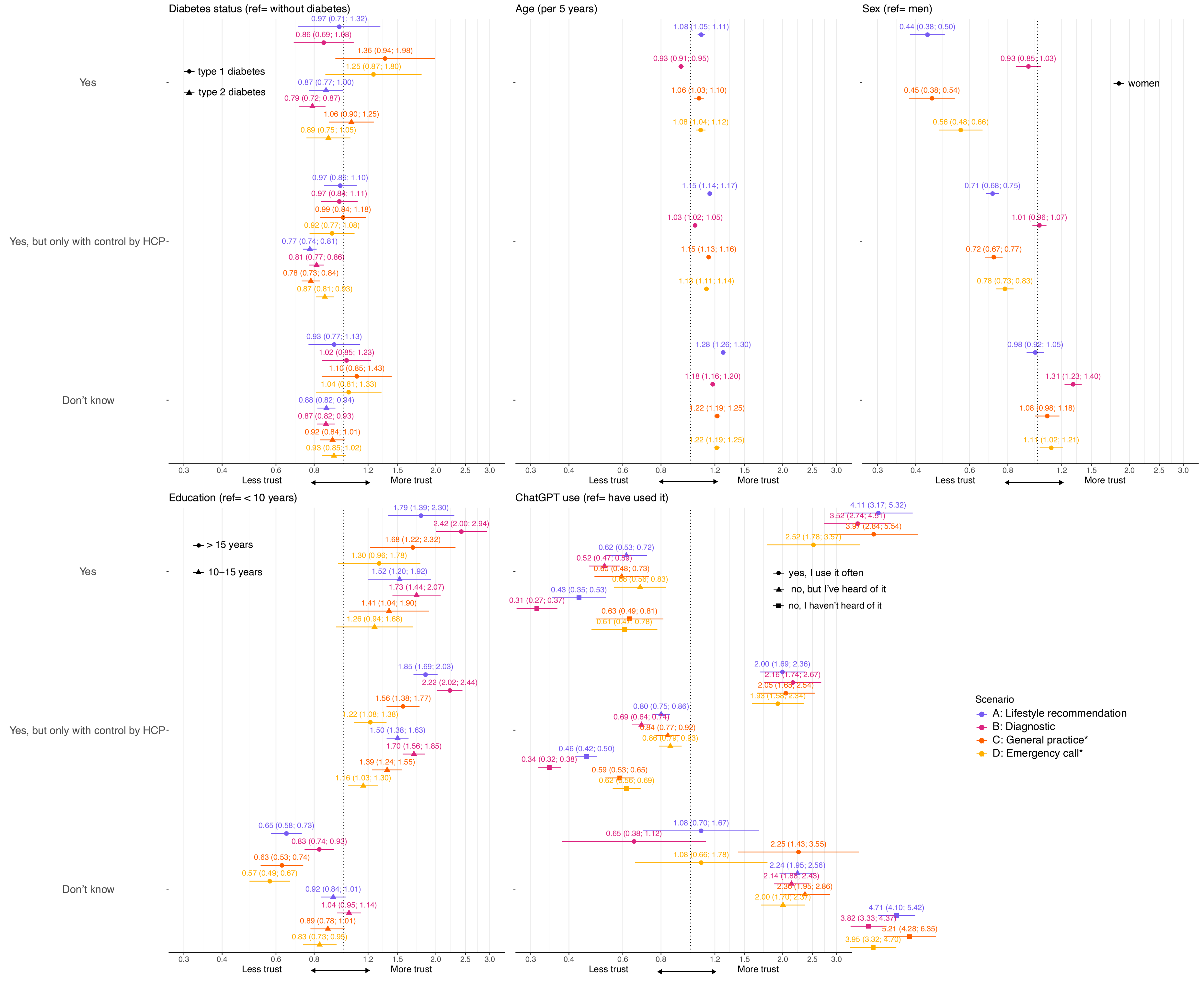
Diabetes status (ref= without diabetes)
type 1 diabetes
type 2 diabetes
Yes
0.97 (0.71; 1.32)
0.86 (0.69; 1.08)
1.36 (0.94; 1.98)
1.25 (0.87; 1.80)
0.87 (0.77; 1.00)
0.79 (0.72; 0.87)
1.06 (0.90; 1.25)
0.89 (0.75; 1.05)
Yes, but only with control by HCP
0.97 (0.86; 1.10)
0.97 (0.84; 1.11)
0.99 (0.84; 1.18)
0.92 (0.77; 1.08)
0.77 (0.74; 0.81)
0.81 (0.77; 0.86)
0.78 (0.73; 0.84)
0.87 (0.81; 0.93)
Don't know
0.93 (0.77; 1.13)
1.02 (0.85; 1.23)
1.10 (0.85; 1.43)
1.04 (0.81; 1.33)
0.88 (0.82; 0.94)
0.87 (0.82; 0.93)
0.92 (0.84; 1.01)
0.93 (0.85; 1.02)
Age (per 5 years)
1.08 (1.05; 1.11)
0.93 (0.91; 0.95)
1.06 (1.03; 1.10)
1.08 (1.04; 1.12)
1.15 (1.14; 1.17)
1.03 (1.02; 1.05)
1.15 (1.13; 1.16)
1.13 (1.11; 1.14)
1.28 (1.26; 1.30)
1.18 (1.16; 1.20)
1.22 (1.19; 1.25)
1.22 (1.19; 1.25)
Sex (ref= men)
women
0.44 (0.38; 0.50)
0.93 (0.85; 1.03)
0.45 (0.38; 0.54)
0.56 (0.48; 0.66)
0.71 (0.68; 0.75)
1.01 (0.96; 1.07)
0.72 (0.67; 0.77)
0.78 (0.73; 0.83)
0.98 (0.92; 1.05)
1.31 (1.23; 1.40)
1.08 (0.98; 1.18)
1.11 (1.02; 1.21)
0.3 0.4 0.6 0.8 1.2 1.5 2.0 2.5 3.0
Less trust
More trust
Education (ref= < 10 years)
> 15 years
10–15 years
1.79 (1.39; 2.30)
2.42 (2.00; 2.94)
1.68 (1.22; 2.32)
1.30 (0.96; 1.78)
1.52 (1.20; 1.92)
1.73 (1.44; 2.07)
1.41 (1.04; 1.90)
1.26 (0.94; 1.68)
1.85 (1.69; 2.03)
2.22 (2.02; 2.44)
1.56 (1.38; 1.77)
1.22 (1.08; 1.38)
1.50 (1.38; 1.63)
1.70 (1.56; 1.85)
1.39 (1.24; 1.55)
1.16 (1.03; 1.30)
0.65 (0.58; 0.73)
0.83 (0.74; 0.93)
0.63 (0.53; 0.74)
0.57 (0.49; 0.67)
0.92 (0.84; 1.01)
1.04 (0.95; 1.14)
0.89 (0.78; 1.01)
0.83 (0.73; 0.95)
ChatGPT use (ref= have used it)
yes, I use it often
no, but I've heard of it
no, I haven't heard of it
4.11 (3.17; 5.32)
3.52 (2.74; 4.51)
3.97 (2.84; 5.54)
2.52 (1.78; 3.57)
0.62 (0.53; 0.72)
0.52 (0.47; 0.59)
0.60 (0.48; 0.73)
0.68 (0.56; 0.83)
0.43 (0.35; 0.53)
0.31 (0.27; 0.37)
0.63 (0.49; 0.81)
0.61 (0.47; 0.78)
2.00 (1.69; 2.36)
2.16 (1.74; 2.67)
2.05 (1.65; 2.54)
1.93 (1.58; 2.34)
0.80 (0.75; 0.86)
0.69 (0.64; 0.74)
0.84 (0.77; 0.92)
0.86 (0.79; 0.93)
0.46 (0.42; 0.50)
0.34 (0.32; 0.38)
0.59 (0.53; 0.65)
0.62 (0.56; 0.69)
1.08 (0.70; 1.67)
0.65 (0.38; 1.12)
2.25 (1.43; 3.55)
1.08 (0.66; 1.78)
2.24 (1.95; 2.56)
2.14 (1.88; 2.43)
2.36 (1.95; 2.86)
2.00 (1.70; 2.37)
4.71 (4.10; 5.42)
3.82 (3.33; 4.37)
5.21 (4.28; 6.35)
3.95 (3.32; 4.70)
Scenario
A: Lifestyle recommendation
B: Diagnostic
C: General practice*
D: Emergency call*

**Figure S4:** Associations between participants characteristics and a positive response to chatbot-based technologies (reference: Yes, with HCP) in different scenarios in the healthcare setting. All estimates are adjusted for use of ChatGPT.

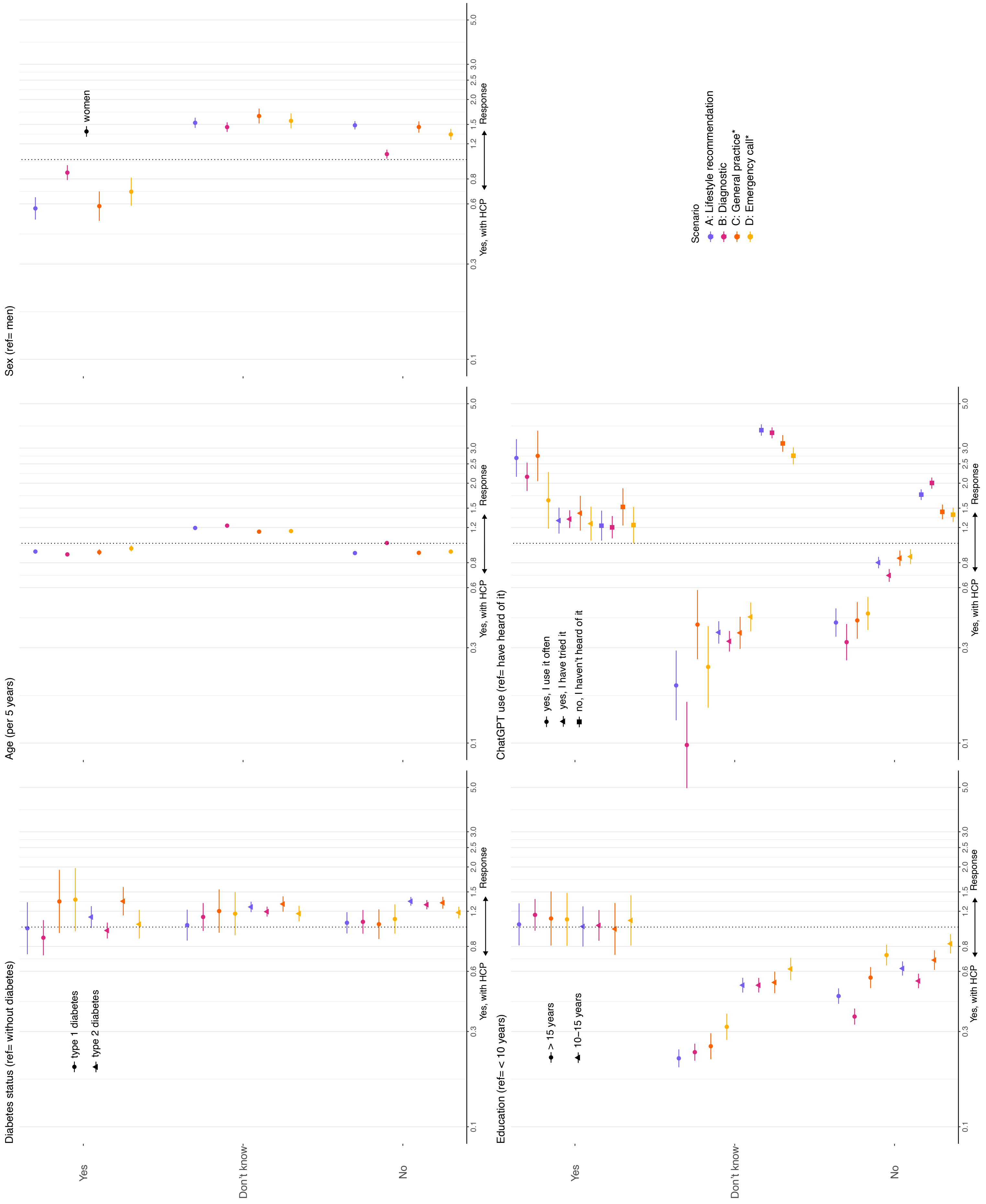

**Figure S5:** Participants' views on the use of chatbot-based solutions in different scenarios in healthcare by diabetes status. The answer addresses to which degree the participant 'would trust the chatbot'.

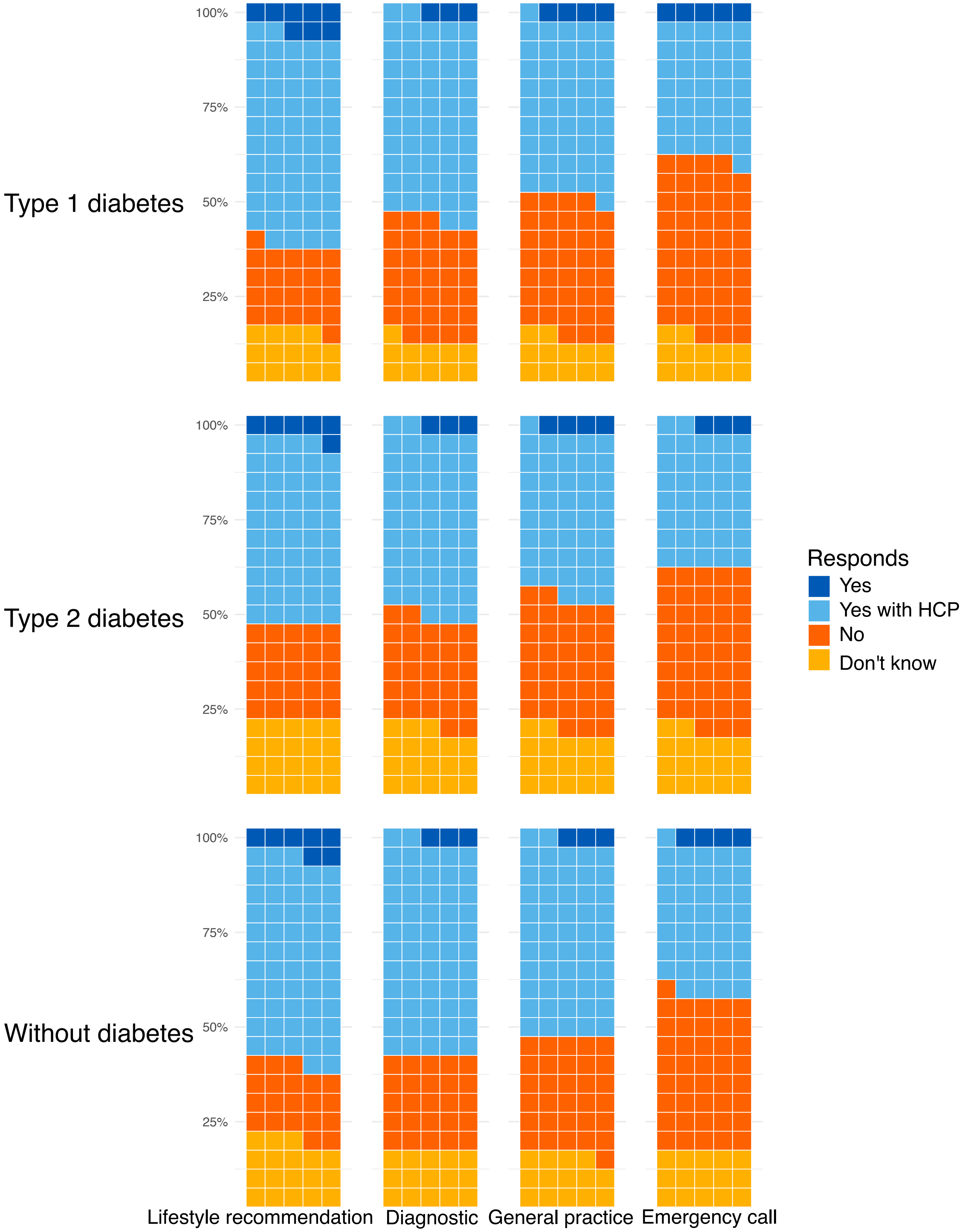

Type 1 diabetes
Type 2 diabetes
Without diabetes
100%
75%
50%
25%
Lifestyle recommendation
Diagnostic
General practice
Emergency call
Responds
Yes
Yes with HCP
No
Don't know
